# A Scalable Framework for Harmonized mtDNA Analysis Across Diverse Biobanks

**DOI:** 10.64898/2026.08.21.26361041

**Authors:** Daniel R. Schecter, Simon SzeKing Lee, Tushar Vimal, Yash Lahoti, Vanessa F. Gonçalves, Kayla Retallick-Townsley, Jiuhong Pang, Aysegul Guvenek, Michael Preuss, Rory J. Tinker, Eva Morava, Tamas Kozicz, Michio Hirano, Jaya Ganesh, Ali Naini, Jingjing Liang, Lea K Davis

**Affiliations:** Department of Genetics and Genomic Sciences, Icahn School of Medicine at Mount Sinai, New York, NY 10029, USA; Center for AI in Children’s Health, Mindich Child Health and Development Institute, Icahn School of Medicine at Mount Sinai, New York, NY 10029, USA; Department of Artificial Intelligence and Human Health, Icahn School of Medicine at Mount Sinai, New York, NY 10029, USA; Department of Pharmacy Practice and Science, University of Arizona, Tucson, AZ 85721, USA; Campbell Family Mental Health Research Institute, Centre for Addiction and Mental Health (CAMH), Toronto, ON, Canada; Tanenbaum Centre for Pharmacogenetics, Campbell Family Mental Health Research Institute, Centre for Addiction and Mental Health, Toronto, ON, Canada; Department of Psychiatry, University of Saskatchewan, Saskatoon, SK, Canada; Charles Bronfman Institute for Personalized Medicine, Icahn School of Medicine at Mount Sinai, New York, NY 10029, USA; Department of Pathology and Cell Biology, Columbia University Irving Medical Center, New York, NY 10032, USA; Department of Neurology, Columbia University Irving Medical Center, New York, NY 10032, USA

**Keywords:** mitochondrial DNA, genomic biobank, whole-exome sequencing, whole-genome sequencing, mtDNA-Server 2, population genomics

## Abstract

Mitochondrial DNA (mtDNA) is increasingly recognized as an important contributor to human disease and population variation, yet most genomic biobanks do not provide standardized mtDNA variant datasets despite abundant mitochondrial sequencing reads in existing whole-exome and whole-genome sequencing data. We developed a scalable framework based on the Mitoverse mtDNA-Server 2 Fusion workflow to generate harmonized, analysis-ready mtDNA resources across diverse biobank infrastructures. The framework was implemented in the Mount Sinai Million Health Discoveries Program (54,151 participants) using the native Nextflow workflow and adapted for the All of Us Research Program (197,361 participants) using a custom cloud implementation that preserved the same analytical strategy. Across 251,512 participants, the framework generated standardized mtDNA datasets containing 12.9 million variant observations suitable for downstream genomic and electronic health record–linked analyses. This framework enables reproducible, population-scale mitochondrial genomics across institutional and national biobanks without requiring additional sequencing or development of new variant-calling methods.

## Introduction

Large genomic biobanks have transformed translational research and human genetics by enabling population-scale analyses of genomic variation linked to longitudinal clinical data. Although whole-exome (WES) and whole-genome (WGS) sequencing datasets generated by large genomic biobanks contain abundant mitochondrial sequencing reads, many resources do not release processed mitochondrial DNA (mtDNA) variant datasets as part of their standard genomic data products(1). As a result, mitochondrial genomic analyses remain inaccessible compared to nuclear genomic data despite the availability of the underlying sequencing data, limiting the scientific and clinical value of existing genomic resources.

Unlike the nuclear genome, mtDNA is maternally inherited and consists of a 16,569-bp circular genome encoding 13 protein-coding genes, 22 transfer RNA (tRNA) genes, and 2 ribosomal RNA (rRNA) genes. Hundreds to thousands of mtDNA molecules are present within each cell, and these copies may differ in sequence within the same individual, a phenomenon known as heteroplasmy (2). Together with mtDNA copy number and mitochondrial haplogroup, these unique features of mitochondrial genetics have increasingly been associated with aging, cancer, cardiometabolic disease, mortality, and primary mitochondrial disease (3,4).

Large sequencing cohorts have made this variation measurable across whole populations. Deep resequencing shows that heteroplasmy is common in healthy individuals and that pathogenic mtDNA variants are observed in the general population more frequently than expected based on the prevalence of clinically manifested mitochondrial disease, highlighting the incomplete penetrance of many mtDNA variants (5–7). Biobank studies of hundreds of thousands of participants have identified nuclear loci that influence heteroplasmy and copy number and have cataloged mtDNA genotype–phenotype associations (8,9). Once high-quality mtDNA variants are generated at the cohort level, they can be analyzed with established genomic and statistical methods, including phenome-wide association studies and integration with longitudinal electronic health record data (EHR) (10,11). Generating accessible mtDNA datasets from existing biobanks has the potential to advance population mitochondrial genomics, variant interpretation, and precision medicine without requiring participants to undergo additional sequencing or enrollment.

Despite this relevance, mtDNA is routinely omitted from the primary analytic products of the resources best positioned to use it. GnomAD, a widely used public catalog of human allele frequencies, excluded mtDNA from its reference until a dedicated pipeline was applied to 56,434 genomes (1). This example illustrates a broader limitation: even when mitochondrial reads are present in the underlying sequencing data, mtDNA variants may remain absent from the genomic resources provided to investigators. Researchers must therefore independently reconstruct mitochondrial datasets, often without standardized workflows, or are unable to incorporate mtDNA into their analyses altogether.

The barrier is computational rather than biological. mtDNA is difficult to call with standard nuclear pipelines: the genome is circular and present at variable copy number, nuclear mitochondrial DNA segments (NUMTs) generate false-positive calls, and the clinically and biologically relevant variation frequently occurs at low heteroplasmy levels that conventional germline variant callers are not designed to detect (1,12). Dedicated tools have substantially improved mtDNA variant detection, including automated heteroplasmy annotation, contamination detection, and NUMT-aware calling (13–15). However, the availability of dedicated mtDNA callers has not by itself resolved the implementation challenge. These tools remain fragmented and adopt different trade-offs between sensitivity and contamination control; their detection thresholds and reference panels vary across platforms and ancestries (16–18). Even where mtDNA reads exist, they are seldom released as an annotated, cohort-level dataset ready for analysis. This gap is largest for ancestrally diverse national biobanks, which remain underrepresented in existing mtDNA reference panels (16,19).

Collectively, existing software has largely solved the problem of mtDNA variant detection. The remaining challenge is implementation: developing a reproducible and scalable framework that can be deployed across diverse biobanks to generate standardized, quality-controlled mtDNA datasets from sequencing resources in which such datasets are not routinely provided. Such a framework must preserve a consistent analytic approach while accommodating differences in sequencing modality, cohort scale, computational environment, and data-access model.

Here, we demonstrate this framework across two complementary genomic resources that differ in scale and computational infrastructure: a large institutional biobank, the Mount Sinai Million Health Discoveries Program (MSM), comprising 54,151 participants; and a national genomic program, the All of Us Research Program (AoU), comprising 197,361 participants included in the present analysis (20). These cohorts provide complementary environments that differ substantially in scale, computational infrastructure, and data-access policies, enabling evaluation of the portability and reproducibility of a common mtDNA analysis framework. These cohorts serve as distinct implementation settings through which to evaluate whether a common mtDNA analytic strategy can be deployed across institutional and national resources.

We processed both cohorts using a single workflow based on the mtDNA-Server 2 Fusion approach, which combines GATK Mutect2 with the mitochondria-specific caller mutserve2 and adds contamination detection and haplogroup assignment (14,21,22). Within the institutional high-performance computing environment, the established containerized workflow could be implemented directly. Because AoU Researcher Workbench does not support unrestricted execution of external containerized workflows, we recreated the core analytical workflow under different computational constraints so that the outputs remained standardized across cohorts. The resulting framework converts previously inaccessible mitochondrial sequencing reads into harmonized variant calls with heteroplasmy level, haplogroup assignments, and quality-control metrics suitable for downstream genomic and EHR–linked analyses.

Rather than introducing a new mtDNA variant caller, this work demonstrates how existing methods can be implemented and adapted to operate reproducibly, portably, and at scale to unlock mtDNA data from genomic resources in which mitochondrial variation was not previously available for research. This approach can support broader implementation of mtDNA analysis across biobanks and accelerate translational research in mitochondrial population genomics, primary mitochondrial disease, and precision medicine.

## Methods

### Overall mtDNA Analysis Framework

A harmonized analytical framework was developed to generate standardized mtDNA variant datasets from existing WES and WGS data (Figure 1). The framework was based on the Mitoverse mtDNA-Server 2 Fusion workflow and consisted of mitochondrial read extraction, independent variant calling using GATK Mutect2 and mutserve2, variant filtering and normalization, integration of variant calls into a unified call set, mitochondrial-specific annotation, contamination assessment, and haplogroup assignment.

**Figure 1.**
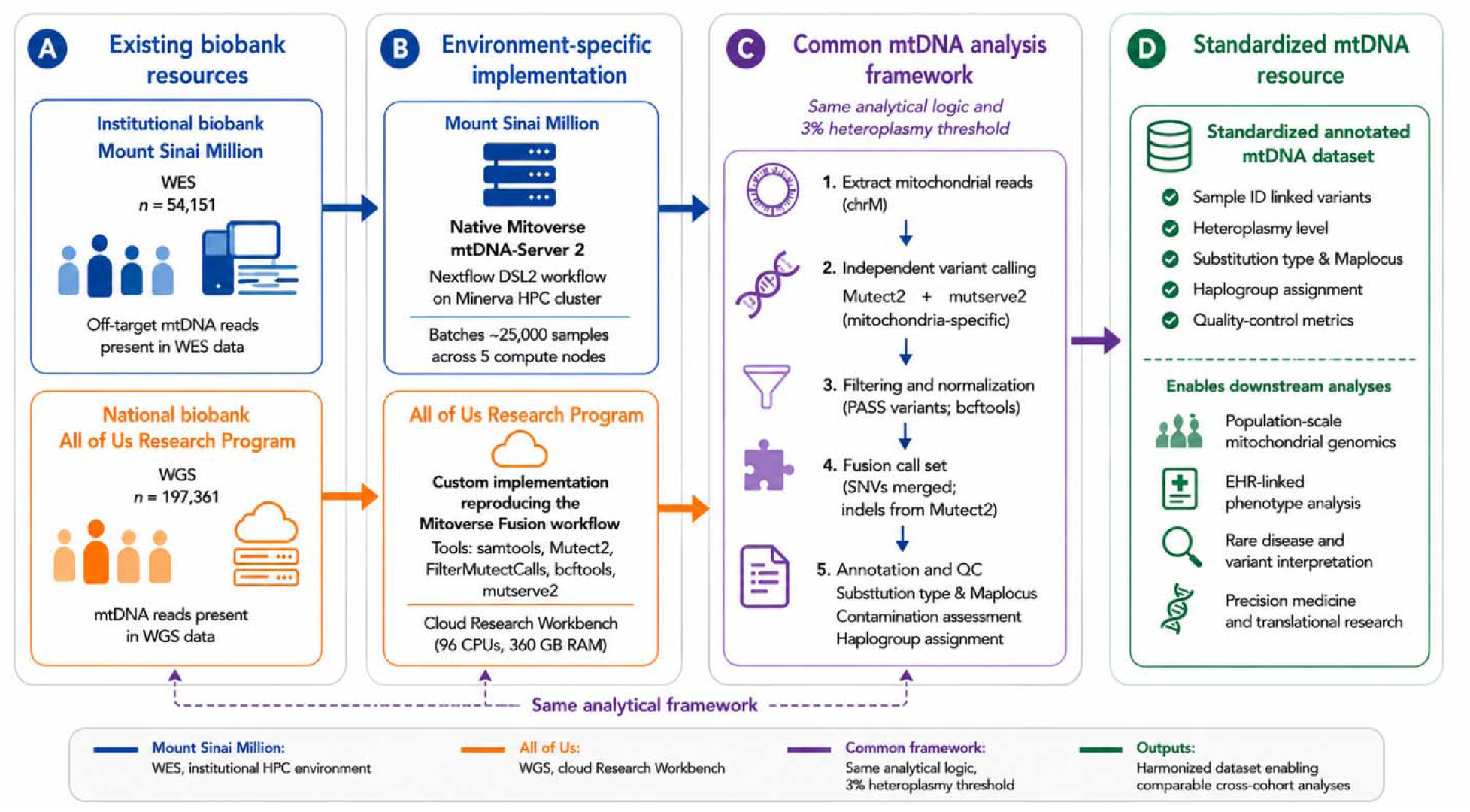
Implementation of the mtDNA analysis framework across genomic biobanks. (A) Cohort inputs. (B) Environment-specific implementation using the native Mitoverse mtDNA-Server 2 Fusion workflow on the Mount Sinai high-performance computing cluster and a custom Fusion-based implementation reproducing the Mitoverse Fusion workflow within the AoU Researcher Workbench. (C) Shared mtDNA analytical workflow consisting of mitochondrial read extraction, independent variant calling with GATK Mutect2 and mutserve2 (3% heteroplasmy threshold), filtering, normalization, Fusion call-set generation, and annotation. (D) Standardized annotated mtDNA dataset containing sample-linked variants, heteroplasmy levels, substitution type and map locus, haplogroup assignments, and quality-control metrics for downstream genomic and EHR-linked analyses.

For the MSM, the framework was implemented directly as a containerized Nextflow DSL2 workflow on the Minerva high-performance computing cluster. For AoU, the framework was adapted for implementation within the Researcher Workbench cloud environment, as described below.

#### 1. Implementation in the Mount Sinai Million

##### **a.** Cohort and pipeline implementation

The MSM is a large-scale institutional genomic biobank led by the Icahn School of Medicine at Mount Sinai in collaboration with the Regeneron Genetics Center. The program integrates genomic sequencing data with de-identified EHRs from a large, ancestrally diverse patient population within the Mount Sinai Health System.

Genetic ancestries of MSM participants were inferred using HapMap3 populations as a reference. Principal components (PCs) were calculated from HapMap3 samples using common SNPs with a minor allele frequency (MAF) > 5%, and MSM samples were projected onto these PCs (23). Kernel density estimates (KDEs) were then trained for each ancestral group using the first four PCs. Based on the KDE results, each MSM sample was assigned to one of five superpopulations according to its genetic similarity to the HapMap3 reference populations: African (AFR, n = 15,644), Admixed American (AMR, n = 8,096), East Asian (EAS, n = 2,535), European (EUR, n = 21,059), or South Asian (SAS, n = 2,312). An additional 4,505 participants had unknown or no available ancestry assignment.

WES data from 54,151 MSM participants aligned to the GRCh38 reference genome were analyzed using the Mitoverse mtDNA-Server 2 bioinformatics pipeline implemented as a Nextflow DSL2 workflow and executed on the Minerva high-performance computing cluster. Mitochondrial variants were identified from off-target mitochondrial sequencing reads obtained during WES. Samples were processed in batches of approximately 4,300 participants across five compute nodes on the Minerva cluster, and batch-level outputs were merged following annotation.

The pipeline was run in Fusion mode, which combines variant calls generated by Mutect2 and mutserve2 into a unified variant call set. In addition to variant calling, the workflow performs quality control, contamination detection, variant annotation, and mitochondrial haplogroup assignment from off-target mitochondrial sequencing reads obtained during WES. Variants were reported using a minimum heteroplasmy detection threshold of 3% (detection_limit = 0.03).

##### **b.** Outputs and cohort integration

For each sample, the pipeline generated the standard Mitoverse annotated tab-delimited text (.txt) output, including genomic coordinates, reference and alternate alleles, heteroplasmy level, sequencing depth, genotype, base quality, variant classification (SNV/indel), predicted functional consequence, amino acid changes, haplogroup assignment, population frequencies, sequencing quality metrics, and mitochondrial-specific annotations from integrated reference databases.

These text files were used for downstream variant interpretation, quality assessment, and genotype–phenotype analyses. Following annotation, sample outputs from each processing batch were merged to create a cohort-level mitochondrial variant dataset. The Nextflow workflow can be rerun as additional MSM sequencing batches become available, allowing new samples to be processed and integrated into the existing mitochondrial variant dataset.

#### 2. Implementation in the All of Us Research Program

##### **a.** Cohort and workflow adaptation

The All of Us (AoU) Research Program is a national precision medicine initiative that integrates whole-genome sequencing, electronic health records, physical measurements, and participant survey data from a diverse U.S. population (24). Analyses were performed using the Controlled Tier v8 (CDR v8) dataset available through the AoU Researcher Workbench.

To evaluate implementation at national biobank scale, we analyzed 197,361 participants from the All of Us Research Program. A large, phenotypically diverse subset of participants was selected using SNOMED CT diagnoses spanning neurologic, cardiovascular, endocrine, renal, audiologic, and ophthalmologic conditions. Mitochondrial DNA analysis was performed in three largest genetically inferred ancestry groups, comprising 125,845 European (EUR), 33,166 Admixed American (AMR), and 38,350 African (AFR) participants (197,361 total). Genetic ancestry assignments were generated by the AoU Research Program using principal component analysis and a classifier trained on Human Genome Diversity Project (HGDP) and 1000 Genomes Project reference populations (25,26).

The Mitoverse mtDNA-Server 2 workflow could not be executed within the AoU Researcher Workbench due to restrictions on Nextflow/Docker/Singularity workflows. We therefore developed a custom implementation to recreate the core analytical workflow of the Mitoverse Fusion pipeline within the AoU environment, while generating standardized outputs compatible with the Mitoverse mtDNA-Server 2 annotation schema. Consistent with the Fusion approach, the custom implementation used both Mutect2 and mutserve2 as variant callers and retained the same 3% heteroplasmy threshold, with caller outputs integrated using the custom workflow described below.

The pipeline was implemented using samtools, GATK Mutect2, FilterMutectCalls, bcftools, and mutserve2. Mutserve2 used the revised Cambridge Reference Sequence (rCRS), whereas Mutect2 variant calls were generated relative to the chrM reference used in GRCh38. Subsequent normalization ensured consistent variant representation across callers.

##### **b.** Pipeline implementation

Pipeline execution was optimized for the AoU Workbench cloud environment using 96 CPUs and 360 GB RAM, with samples processed in batches of approximately 20,000 participants.

For each participant, the CRAM file and corresponding index file were retrieved from the AoU workspace. Chromosome M (chrM) reads were streamed directly from the CRAM file using samtools to generate a chrM BAM file. Consistent with the Mitoverse Fusion approach, the BAM file was analyzed independently using both Mutect2 and mutserve2. Mutect2 variant calls were filtered using FilterMutectCalls, while mutserve2 was run using a 3% heteroplasmy detection threshold.

The outputs from both callers were normalized against the chrM.fa reference genome using bcftools to ensure consistent variant representation. Only variants marked PASS were retained. Because mutserve2 reports only SNVs, indels were obtained exclusively from the Mutect2 output. The Mutect2 VCF headers were standardized to match the mutserve2 format, after which PASS SNVs from both mutserve2 and Mutect2 were merged and redundant SNVs were removed. The resulting SNV set was then combined with Mutect2-derived indels to generate a unified mtDNA VCF for each participant.

##### **c.** Standardized outputs

The pipeline generated two output files for each sample: (1) a unified mtDNA VCF and (2) an annotated tab-delimited text (.txt) file compatible with the standard Mitoverse mtDNA-Server 2 output schema. The standardized text files were used for downstream variant interpretation, quality assessment, and genotype–phenotype analyses alongside the MSM data.

## Results

### Biobank-scale processing performance

The complete MSM cohort of 54,151 participants was processed in six batches of approximately 4,326 participants using 8 CPU cores and 64 GB memory, requiring approximately 112 hours of total runtime. Within the AoU Workbench, batches of approximately 20,000 participants were processed using 96 CPUs and 360 GB RAM. Each batch required approximately 60–72 hours of runtime and incurred a computational cost of approximately US$300 (Table 1). All processing batches completed successfully, generating standardized participant-level mtDNA variant datasets for all 251,512 participants included in the analysis

**Table 1.**
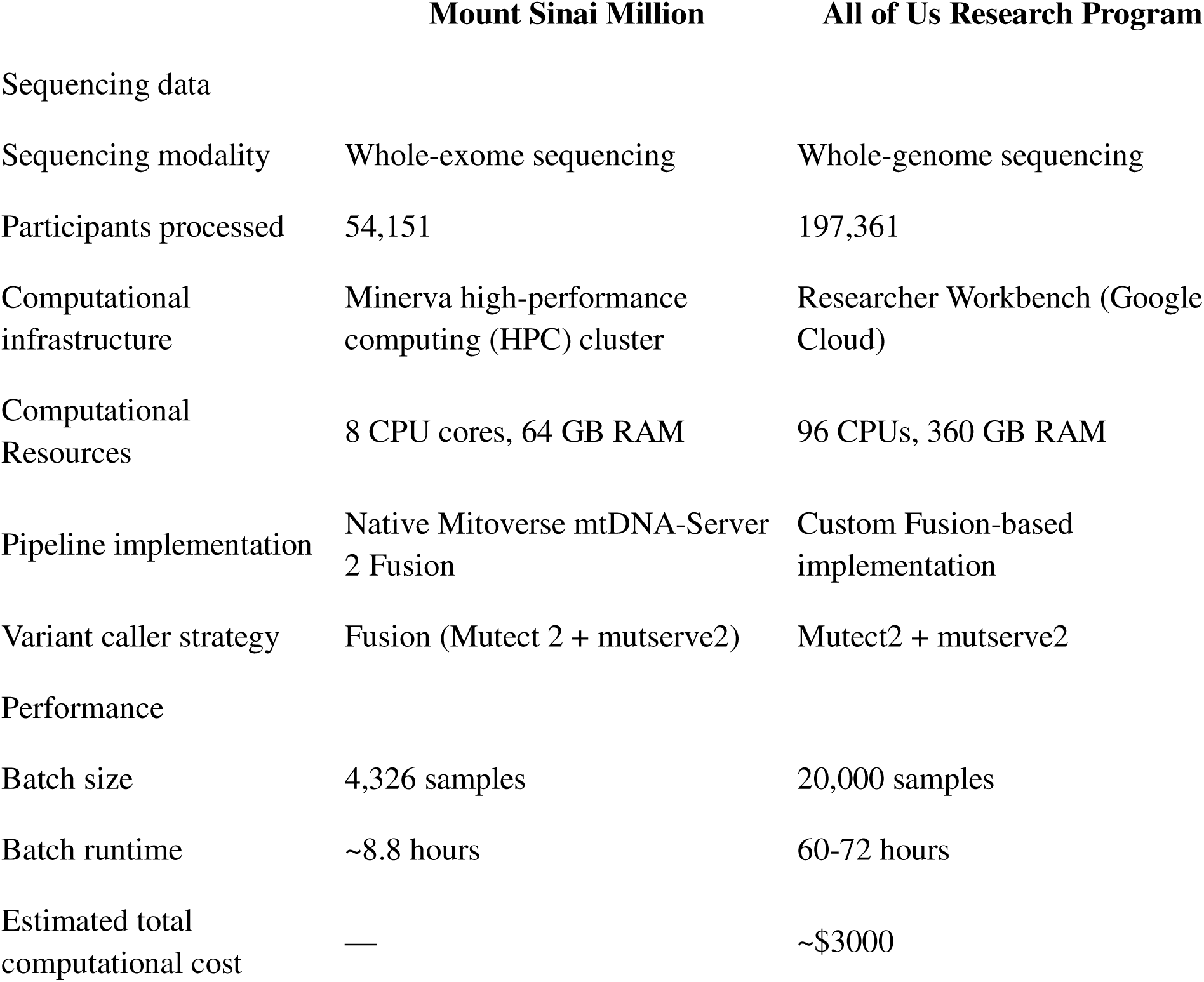
Implementation of the harmonized mtDNA analysis framework across two genomic biobanks. Comparison of the implementation of the harmonized mtDNA analysis framework in the MSM and the AoU Research Program. The table summarizes differences in sequencing modality, cohort size, computational infrastructure, pipeline implementation, batch processing, runtime, and computational cost while demonstrating a common Fusion-based analytical strategy that generated standardized participant-level mtDNA outputs across both biobanks.

|  | Mount Sinai Million | All of Us Research Program |
| --- | --- | --- |
| Sequencing data |  |  |
| Sequencing modality | Whole-exome sequencing | Whole-genome sequencing |
| Participants processed | 54,151 | 197,361 |
| Computational infrastructure | Minerva high-performance computing (HPC) cluster | Researcher Workbench (Google Cloud) |
| Computational Resources | 8 CPU cores, 64 GB RAM | 96 CPUs, 360 GB RAM |
| Pipeline implementation | Native Mitoverse mtDNA-Server 2 Fusion | Custom Fusion-based implementation |
| Variant caller strategy | Fusion (Mutect 2 + mutserve2) | Mutect2 + mutserve2 |
| Performance |  |  |
| Batch size | 4,326 samples | 20,000 samples |
| Batch runtime | ~8.8 hours | 60-72 hours |
| Estimated total computational cost | — | ~\$3000 |

### Generation of mtDNA resources

The MSM implementation generated 3,497,139 mtDNA variant observations from 54,151 participants, representing 16,128 unique mitochondrial variants. Participants had a mean of 64.6 variant observations and a median of 58 variant observations per participant (interquartile range [IQR], 37–84).

The AoU implementation generated 9,461,434 mtDNA variant observations from 197,361 participants, representing 34,142 unique mitochondrial variants. Participants had a mean of 47.9 variant observations and a median of 45 variant observations per participant (IQR, 34– 60) (Figure 2).

**Figure 2.**
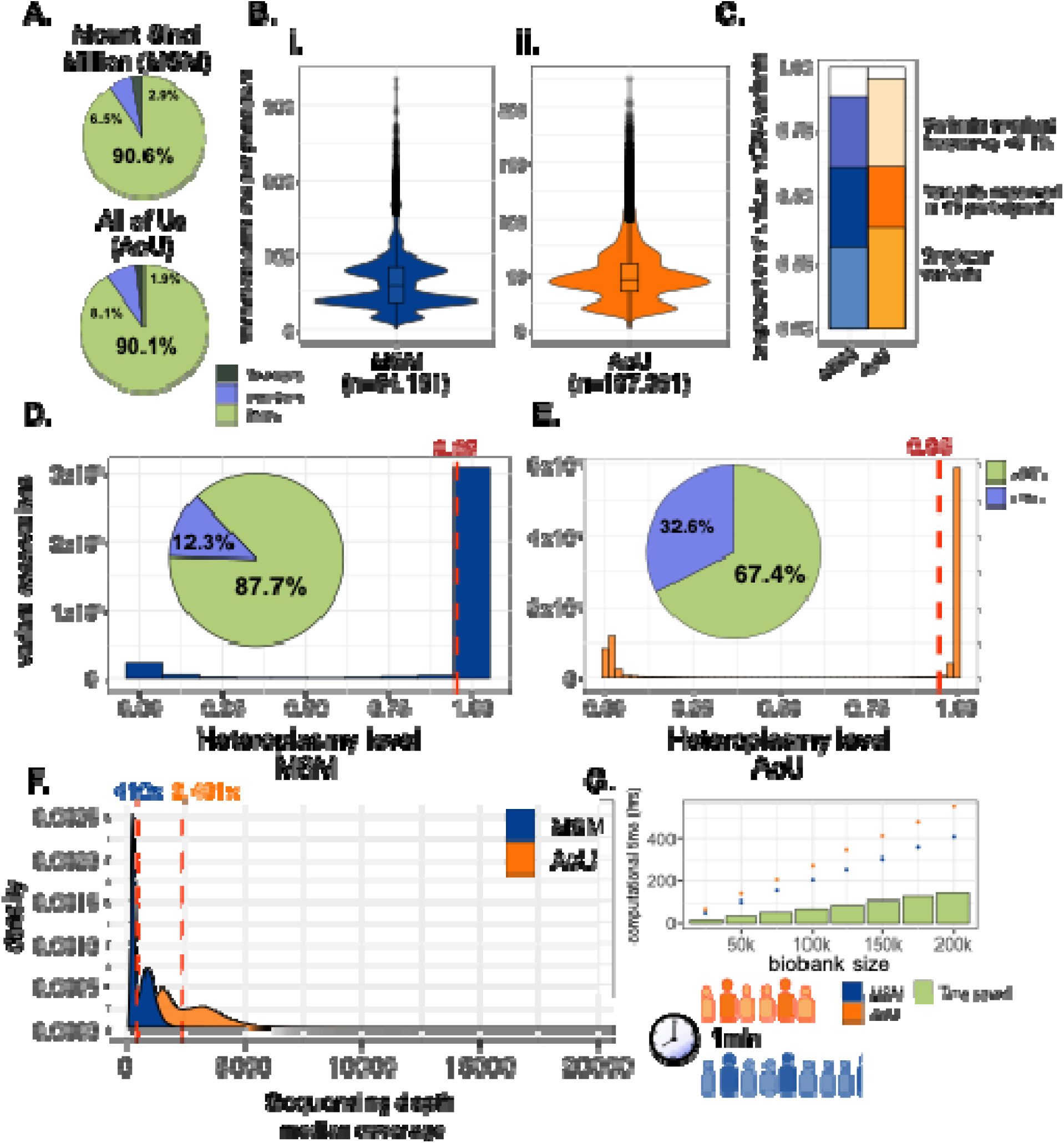
Comparison of mtDNA datasets generated from the MSM and the AoU following implementation of the harmonized workflow. (A) Variant type composition of SNVs, deletions, and insertions in the Mount Sinai Million (MSM; top) and All of Us (AoU; bottom) biobanks. (B) Variant burden per participant in (i) MSM and (ii) AoU. Violin plots show the total variant observations per participant with overlaid boxplots. (C) Cumulative proportion of rare mtDNA variants (bottom bar = singleton variants, middle bar = variants observed in ≤5 participants, and top bar = variants with a cohort frequency <0.1%) in MSM (blue) and AoU (orange). (D–E) Heteroplasmy level distributions in (D) MSM and (E) AoU demonstrate a near-homoplasmic skew; pie charts represent the proportion of variants with >95% heteroplasmy (green). (F) Distribution of median participant mtDNA sequencing depth in MSM (blue; whole-exome sequencing) and AoU (orange; whole-genome sequencing). (G) Estimated computational time (hours) required to analyze biobanks containing 25,000–200,000 participants based on the observed performance of the native Mitoverse mtDNA-Server 2 Fusion workflow in MSM (blue dots) and the custom implementation in AoU (orange dots), with the estimated time saved (green bars) at each population size.

Overall, the harmonized framework generated 12,958,573 mtDNA variant observations from 251,512 participants across the two biobanks. Together, the two implementations produced standardized, analysis-ready mtDNA datasets suitable for downstream genomic and phenotype association analyses.

### Comparison of mtDNA datasets

The characteristics of the generated mtDNA datasets reflected the underlying differences between WES and WGS while demonstrating similar overall variant composition (Table 2; Figure 2). Median participant-level mitochondrial sequencing depth was 410× in MSM (IQR, 219–874×) compared with 2,401× in AoU (IQR, 1,599.5–3,436.5×), consistent with the greater mitochondrial read depth expected from WGS compared with mitochondrial reads obtained from WES (27). In MSM, 3,217,596 of 3,497,139 variant observations (92.0%) passed all quality-control filters. After filtering, all retained variant observations in the AoU Fusion dataset satisfied PASS quality criteria.

**Table 2.** Characteristics of the mtDNA variant datasets generated using the harmonized analysis framework. Comparison of the standardized mtDNA variant datasets generated from the MSM and the AoU following implementation of the harmonized workflow. The table summarizes variant burden, sequencing coverage, quality metrics, variant composition, and rare variant characteristics, demonstrating that the harmonized framework generated comparable, analysis-ready mtDNA resources from both WES and WGS data.

|  | Mount Sinai<br>Million | All of Us<br>Research Program |
| --- | --- | --- |
| Dataset summary |  |  |
| Variant observations | 3,497,139 | 9,461,434 |
| Unique mtDNA variants | 16,128 | 34,142 |
| Variant burden |  |  |
| Mean variants per participant | 64.6 | 47.9 |
| Median variants per participant (IQR) | 58 (37–84) | 45 (34–60) |
| Median participant mtDNA sequencing depth (IQR) | 410× (219–874×) | 2,401× (1,599.5–3,436.5×) |
| PASS variant observations | 3,217,596 (92.0%) | 9,461,434 (100%) |
| Variant composition |  |  |
| SNVs | 3,166,587 (90.6%) | 8,521,635 (90.1%) |
| Insertions | 227,698 (6.5%) | 762,892 (8.1%) |
| Deletions | 102,854 (2.9%) | 176,907 (1.9%) |
| Rare variant characteristics |  |  |
| Singleton variants | 4,969 (30.8%) | 13,159 (38.5%) |
| Variants observed in ≤5 participants | 9,771 (60.6%) | 20,832 (61.0%) |
| Variants with cohort frequency <0.1% | 14,192 (88.0%) | 32,221 (94.4%) |

Single-nucleotide variants comprised the majority of observations in both resources, representing 90.6% of MSM variant observations and 90.1% of AoU observations. Insertions accounted for 6.5% and 8.1% of observations, respectively, while deletions represented 2.9% and 1.9%.

Both implementations captured substantial rare mitochondrial variation (Figure 2C). Approximately 61% of unique mitochondrial variants in both cohorts were observed in five or fewer participants despite differences in sequencing modality, cohort size, ancestry composition, and computational environment (Figure 2). In MSM, 4,969 unique variants (30.8%) were singletons, 9,771 variants (60.6%) were observed in five or fewer participants, and 14,192 variants (88.0%) had a cohort frequency below 0.1%. Similarly, in AoU, 13,159 variants (38.5%) were singletons, 20,832 variants (61.0%) were observed in five or fewer participants, and 32,221 variants (94.4%) had a cohort frequency below 0.1%.

Most detected variant observations were near-homoplasmic (Figure 2D,E). In MSM, 87.7% of observations had heteroplasmy levels greater than 95%, compared with 67.4% of observations in AoU. The difference in heteroplasmy distributions between cohorts may reflect differences in sequencing modality and mitochondrial read depth, including greater sensitivity for detection of low-level heteroplasmy with higher-depth WGS, and should therefore not be interpreted as a biological difference between cohorts.

## Discussion

In this study, we demonstrate that a harmonized mitochondrial DNA analysis framework can be implemented reproducibly across both institutional and national genomic biobanks despite differences in sequencing modality, computational infrastructure, and data-access constraints. Although WES and WGS datasets generated by large genomic biobanks contain abundant mitochondrial sequencing reads, most do not provide processed mtDNA variant datasets comparable to the standardized resources routinely available for the nuclear genome (1). As a result, mitochondrial genomic analyses remain underutilized despite the availability of the underlying sequencing data.

Rather than developing a new variant caller, we show that existing sequencing datasets containing mitochondrial reads can be transformed into standardized, analysis-ready mtDNA resources at population scale. Across 251,512 participants, the framework generated harmonized participant-level mtDNA datasets suitable for downstream genomic analyses and linkage with longitudinal EHR data. While the native and custom implementations both used Mutect2 and mutserve2 and generated standardized output structures, implementation within the AoU environment required adaptation of the workflow, including custom integration of caller outputs. These findings provide a practical and scalable implementation framework that can facilitate broader incorporation of mtDNA into large-scale genomic research.

The generated datasets displayed variant characteristics consistent with previous population-scale mitochondrial sequence studies, including the predominance of near-homoplasmic variants and high proportion of rare mtDNA variation (1). The generation of standardized mtDNA datasets across two biobanks with distinct sequencing modalities and computational environments demonstrates the scalability and portability of the harmonized analytical framework.

A common concern when identifying mitochondrial variants, particularly from off-target reads generated during WES, is the potential for false-positive variant calls arising from nuclear mitochondrial DNA segments (NUMTs) (28). Because NUMTs share extensive sequence homology with the mitochondrial genome, misalignment of nuclear-derived reads may introduce artifactual heteroplasmic variants if not appropriately addressed. Rather than relying on conventional germline variant callers alone, the Mitoverse mtDNA-Server 2 Fusion workflow incorporates several features designed to reduce these artifacts (14,29). Fusion combines variant calls from the general-purpose caller GATK Mutect2 and the mitochondria-specific caller mutserve2. The workflow additionally incorporates contamination assessment, mitochondrial-specific quality control, standardized filtering, and haplogroup assignment before generation of the final call set. The workflow has also demonstrated high performance in a benchmarking study comparing multiple mtDNA variant-calling methods (18,30). Although no bioinformatic workflow can completely eliminate the possibility of NUMT-derived calls, particularly in challenging genomic regions or low-level heteroplasmy, use of an established, validated mitochondrial-specific workflow provides substantially greater confidence than standard nuclear variant calling pipelines for large-scale mtDNA analyses.

Despite providing a reproducible framework for generating standardized mtDNA resources from existing sequencing data, several limitations should be considered. For WES, mitochondrial coverage is derived from off-target reads and may vary depending on the exome capture kit, library preparation, sequencing platform, and protocol. Consequently, combining datasets across institutions or sequencing studies may introduce batch effects and complicate cross-study comparisons or meta-analyses (31,32). In addition, WES has reduced sensitivity for detecting low-level heteroplasmy compared with WGS, and the absence of a universally accepted heteroplasmy threshold may contribute to differences in variant reporting across studies (18,33). Because the MSM and AoU implementations were applied to distinct cohorts generated using different sequencing modalities, no participants were analyzed using both implementations, precluding direct sample-level comparison of variant calls between workflows. Finally, because mtDNA variation is strongly structured by maternal ancestry and mitochondrial haplogroup, population stratification should be considered in downstream analyses. Where appropriate, adjustment for nuclear genetic PCs and careful consideration of haplogroup structure may help reduce confounding in genotype–phenotype association studies (34).

As genomic biobanks continue to expand in size and diversity, standardized mitochondrial analysis frameworks will become increasingly important for integrating mtDNA into population genomics and precision medicine. Our study demonstrates that existing WES and WGS datasets can be systematically transformed into standardized, participant-level mtDNA resources using a reproducible and scalable analytical framework. By enabling consistent implementation across distinct sequencing modalities and computational environments, this framework reduces a practical barrier to incorporating mitochondrial variation into genomic research without requiring additional sequencing. The resulting analysis-ready mtDNA datasets provide a foundation for future genotype-phenotype association studies, multi-institutional research consortia, and future investigations of mitochondrial genetics in both common and rare diseases.

## Data availability

The data for this study was obtained from the Mount Sinai Million Biobank, and AoU, all of which restricts the sharing of genetic and clinical data to approved investigators. Additional Mount Sinai Biobank data are not currently publicly available due to restrictions on the data.

## Acknowledgements

Funding for this study was provided by the American Academy of Pediatrics Resident Research Grant Award (2024), United Mitochondrial Disease Foundation– North American Mitochondrial Disease Consortium Gateway to Mitochondrial Medicine Grant (2024), the Healthy Americas Research Consortium (HARC) Grant 2026 (NIH award OT2OD025277), and Alzheimer’s Association AARG grant 23AARG-1019863. This work was supported in part through the Minerva computational and data resources and staff expertise provided by Scientific Computing and Data at the Icahn School of Medicine at Mount Sinai and supported by the Clinical and Translational Science Awards (CTSA) grant UL1TR004419 from the National Center for Advancing Translational Sciences. We gratefully acknowledge the individuals who’s de-identified clinical and genomic data made this work possible. Although this study was conducted using de-identified data, we recognize that these data represent the health information of individuals.

## Abbreviations

mtDNA: mitochondrial DNA
NUMTs: nuclear mitochondrial DNA segments
WES: whole-exome sequencing
WGS: whole-genome sequencing
EHR: electronic health record
MSM: Mount Sinai Million Health Discoveries Program
AoU: All of Us Research Program
HPC: high-performance computing
GRCh38: Genome Reference Consortium Human Build 38
rCRS: revised Cambridge Reference Sequence
chrM: Chromosome M
tRNA: transfer RNA
rRNA: ribosomal RNA
SNV: single-nucleotide variant
BAM: Binary Alignment/Map
CRAM: Compressed Reference-oriented Alignment Map
VCF: Variant Call Format
PCs: principal components
KDEs: kernel density estimates
MAF: minor allele frequency
HGDP: Human Genome Diversity Project
1KGP: 1000 Genomes Project
AFR: African
AMR: Admixed American
EUR: European
EAS: East Asian
SAS: South Asian
SNOMED CT: Systematized Nomenclature of Medicine Clinical Terms
IQR: interquartile range

